# The changing impact of vaccines in the COVID-19 pandemic

**DOI:** 10.1101/2022.03.10.22272222

**Authors:** Jamie A. Cohen, Robyn M. Stuart, Jasmina Panovska-Griffiths, Edinah Mudimu, Romesh G. Abeysuriya, Cliff C. Kerr, Michael Famulare, Daniel J. Klein

## Abstract

The Omicron wave has left a global imprinting of immunity which changes the COVID landscape. In this study, we simulate six hypothetical variants emerging over the next year and evaluate the impact of existing and improved vaccines. We base our study on South Africa’s infection- and vaccination-derived immunity. Our findings illustrate that variant-chasing vaccines will only add value above existing vaccines in the setting where a variant emerges if we can shorten the window between variant introduction and vaccine deployment to under three weeks, an impossible time-frame without significant NPI use. This strategy may have global utility, depending on the rate of spread from setting to setting. Broadly neutralizing and durable next-generation vaccines could avert over three-times as many deaths from an immune-evading variant compared to existing vaccines. Our results suggest it is crucial to develop next-generation vaccines and redress inequities in vaccine distribution to tackle future emerging variants.

## 1 Introduction

In 2020, the world achieved the fastest vaccine development timeline in history, resulting in multiple highly efficacious vaccines against SARS-CoV-2 available to the public in 11 months. Clinical trials showed that these vaccines dramatically reduce the risk of SARS-CoV-2 infection, mild symptoms, and severe COVID-19 (1; 2; 3; 4). Despite the incredible success of the vaccines, their value is ever-changing as a larger share of the global population has some immunity from either infection and/or vaccination and as the virus evolves to evade immunity (5).

Most COVID-19 vaccines are designed to target the spike protein and stimulate production of neutralizing antibodies that fight infection and CD4+ and CD8+ T-cells, which stop disease progression (6). Neutralizing antibodies wane, contributing to falling vaccine effectiveness against infection over time and against immune evading variants (7). At the same time, memory B- and T-cells are relied upon to stimulate an even broader response when the body is exposed to a new virus. Severe disease protection has been more durable, because the disease course for COVID-19 occurs timescales more amenable to the response time of memory B- and T-cells, and less impacted by viral evolution (8; 9; 10; 11; 12).

The combination of waning immunity and the continued emergence of immune-evading variants has prompted many jurisdictions across the world to deliver booster vaccines, which provide temporarily increased protection against infection and broaden the immune response. Scientists additionally are racing to leverage the mRNA platform to quickly develop variant-targeted vaccines to maximize vaccine effectiveness against immune evading variants. At the same time, antiviral pills that can be taken orally outside of a healthcare setting, such as Molnupiravir and Paxlovid, are showing efficacy against severe outcomes if taken shortly after symptom onset (13). As these treatment options grow and become more globally available, they will undoubtedly play a significant role in the ever-changing COVID-19 landscape.

In this study, we explore the changing value of vaccines in a landscape of dynamic immunity and rapidly evolving variants of concern. We use Covasim, an established agent-based model enhanced with detailed intra-host dynamics, to perform our analyses and base our study on a South Africa-like population, including population size, demographics, and previous infection- and vaccination-derived immunity (14). We evaluate the efficacy of vaccines over time and as a function of population immunity from infection, and then generalize these results to consider the marginal impact of expanded vaccine coverage and ongoing boosters given uncertain and unpredictable future variants. As the marginal impact of vaccines falls, we evaluate the broadening of our prevention tools to reduce the impact of COVID-19.

## 2 Results

### 2.1 vaccine effectiveness over time

We begin by evaluating population-level vaccine effectiveness, as quantified by reduction in the risk of severe disease for vaccinated individuals compared to unvaccinated individuals, as a function of time and incremental to their prior immunity. We find that vaccine effectiveness peaks in the month prior to each emerging wave and declines rapidly thereafter. vaccine effectiveness is highest at the beginning of the pandemic and declines over time as the percentage of the population with immunity from prior infection increases. vaccine effectiveness increases more in advance of a more virulent strain, as seen in the level of protection in advance of Delta, despite 50% prior exposure, compared to a less virulent strain, as seen in the level of protection against Omicron (see Fig. 1).

**Fig. 1.**
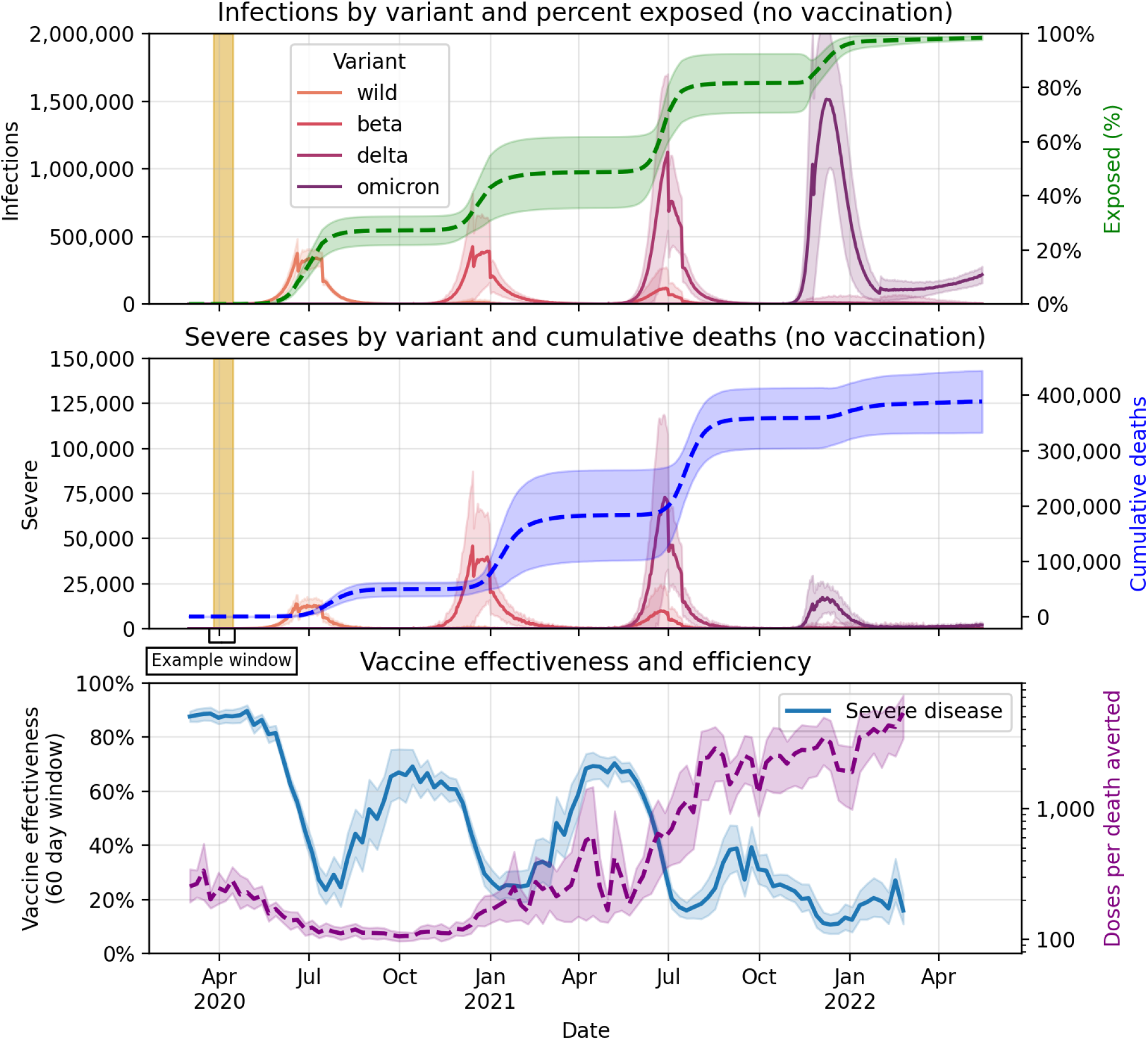
Population-level vaccine effectiveness as a function of time. The top two panels show new infections and new severe cases by variant, percent of population with prior infection, and cumulative deaths in the absence of vaccination. The last panel shows vaccine effectiveness against severe disease over time, which is calculated for the 60 days following vaccine completion (i.e., after second dose) and doses per death averted over time, which is calculated for the cumulative deaths following each vaccine day for the remainder of the simulated period. The golden rectangle represents an example vaccine effectiveness window. Simulations are for a South Africa-like population of 54 million people.

The efficiency of vaccination, as quantified by doses per death averted, is also a function of time, with efficiency decreasing over time at an increasing rate (see Fig. 1). At its peak efficiency, fewer than 100 doses would have been required to avert a single death (or put another way, under 50 people needed to be fully vaccinated). By the end of January 2022, following the Omicron wave, nearly 4,000 doses (or 2,000 people vaccinated) would be required to avert a single death. In comparison, an analysis of childhood and adolescent routine and non-routine immunizations for 10 pathogens estimated an average of 208 fully vaccinated people per death averted over 2000-2019 (15).

### 2.2 Vaccine impact in face of new variants

In the next part, we quantify the dynamic impact of vaccines in the face of uncertain variants of concern. Before considering any additional vaccination, we explore the epidemic dynamics for each of the variants and timelines considered. The first major finding is that the impact of a new variant is more sensitive to the characteristics of the variant (i.e., antigenic distance from sources of prior immunity) than the timeline of introduction (i.e., months after prior Omicron wave). We find a dose-response with the antigenic distance of a new variant and the size of its impact in terms of both new infections and new severe cases (see Fig 2).

**Fig. 2.**
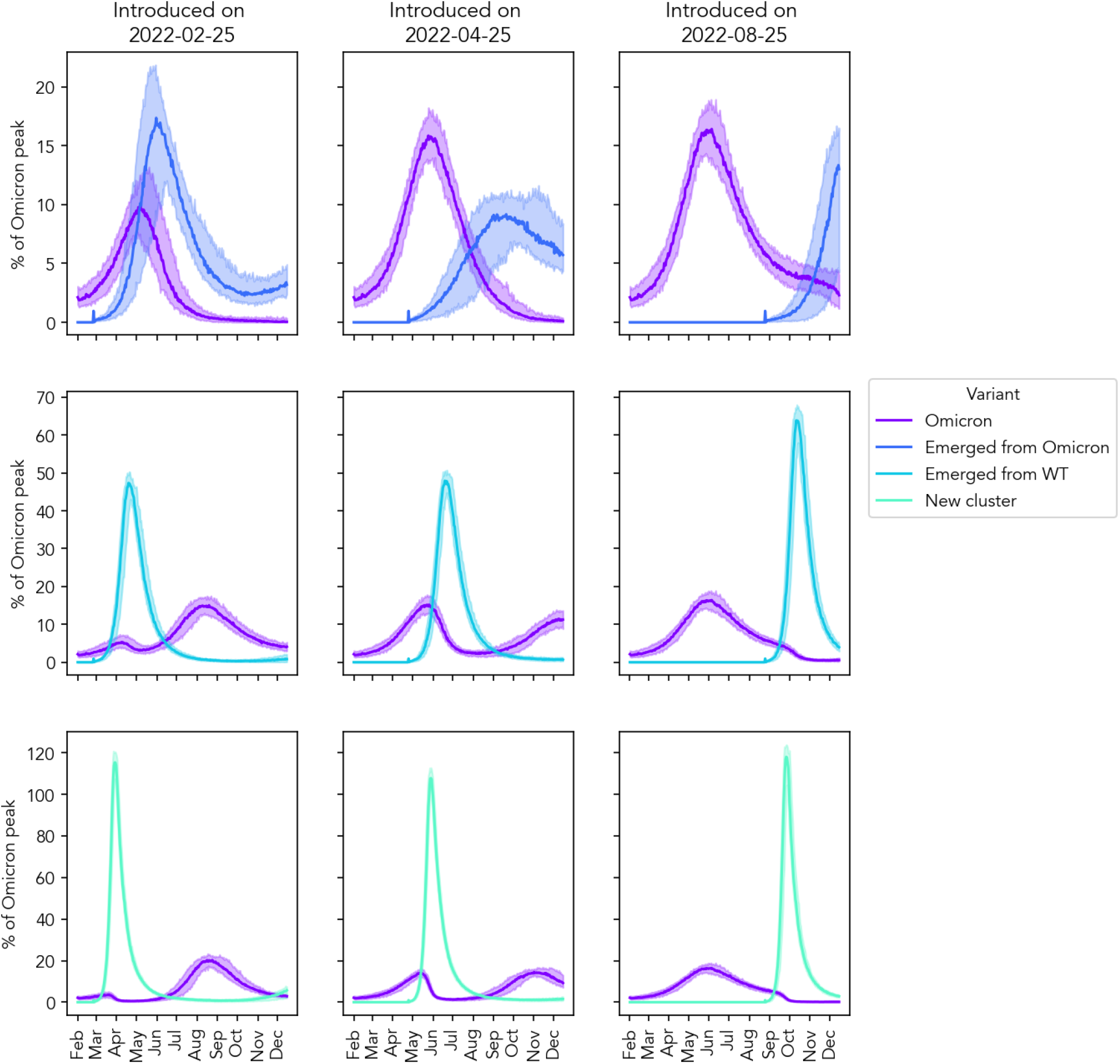
New infections as a percentage of Omicron peak for each of the new variants and variant introduction dates considered. All plots are in the absence of any additional vaccination (‘Status quo’), other non-pharmaceutical interventions, or behavior change.

A variant that emerges from Omicron (i.e., antigenically closest to the most recent variant) would compete directly with a second wave of Omicron. The date of introduction is highly influential in the resulting epidemic dynamics. If this variant emerges within four months after Omicron, it might dampen and even out-compete the re-emergence of Omicron. If the variant emerges later, the second wave of the Omicron would push out the growth of the new variant (see Fig 2).

A variant that emerged from the ancestral cluster or a new cluster entirely (i.e., is antigenically distinct from the most recent variant) would result in a larger growth in cases and its dynamics would be less sensitive to timing of introduction. A variant that emerged from Omicron (i.e., an antigenically similar variant to the most recent variant) would result in a wave of cases 10-20% of the prior wave peak; a variant that emerged from wild-type (i.e., antigenically similar to historical variants) would lead to a wave of cases 50-70% of the prior wave peak; and a variant in a new antigenic cluster entirely would lead to a wave up to 120%.

Variants that have not evolved to become more virulent would result in a similar growth in severe cases, with no obvious further decoupling than has already been seen so far in the pandemic. However, if the variant evolves to become more virulent, there would be an associated increase in severe cases.

### 2.3 Trade-off between primary series and booster dose coverage

In the next part, we quantify the dynamic trade-off between increasing primary series (first and second-dose) and booster-dose coverage of vaccination. We focus first on a relatively well-matched vaccine and variant (proxied here by a variant that emerged from the ancestral strain cluster, which we assume evades 50% of vaccine NAbs), to narrow in on the trade-off between primary series and booster dose coverage, before considering the ongoing impact of antigenic drift. Results indicate that increasing primary series coverage (i.e., decreasing the number of unvaccinated individuals in the population) has a greater marginal and absolute health impact than increasing booster dose coverage among already vaccinated individuals (see Fig 3). Depending upon the date of the introduction of the new variant, a 5-fold increase in primary series coverage would triple the percentage of deaths averted. The same increase in booster dose coverage would double the percentage of deaths averted. Increasing primary dose coverage would achieve 95% of the total possible benefit of increasing coverage among all individuals.

**Fig. 3.**
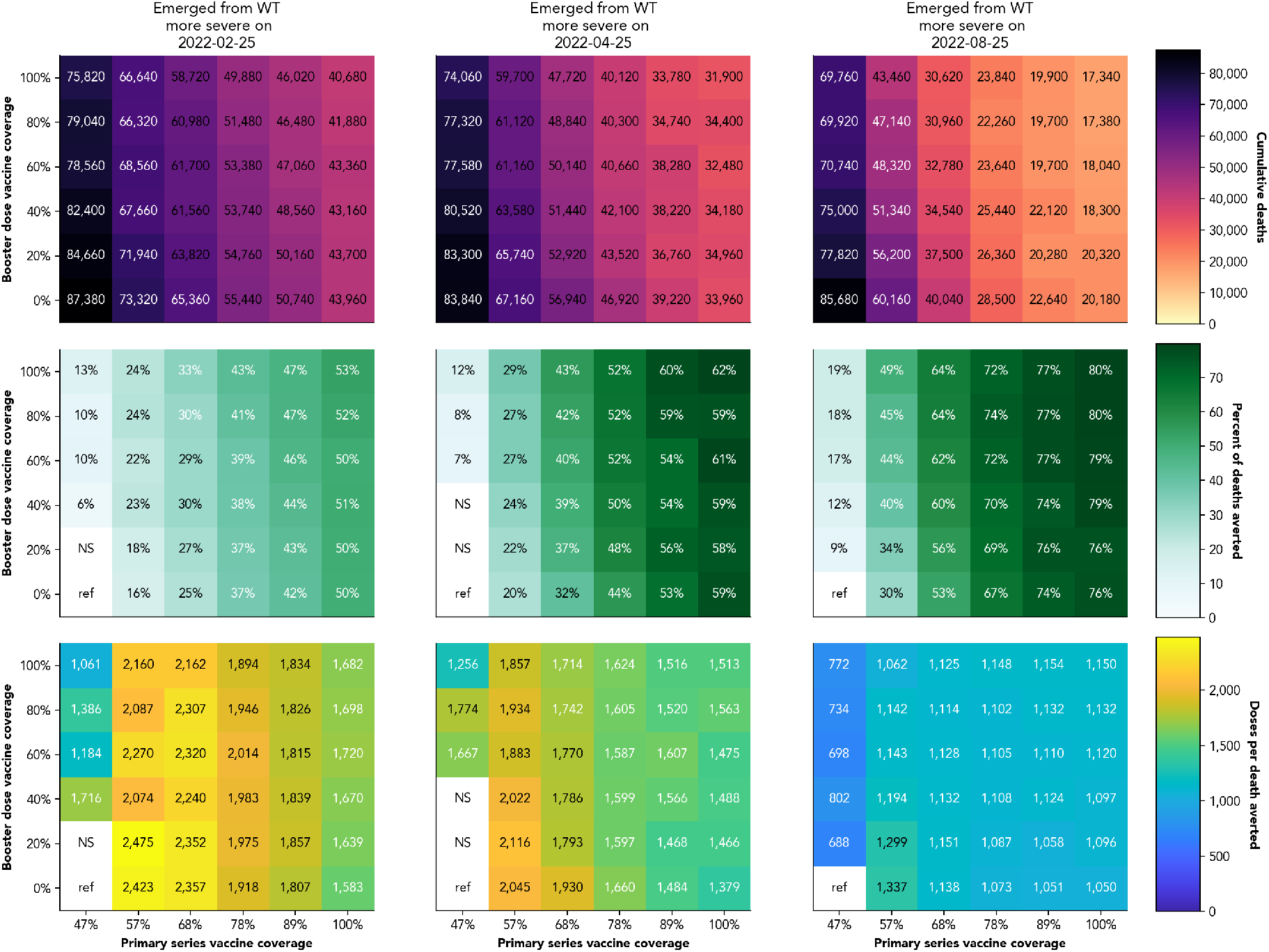
Vaccine impact and efficiency across primary series and booster-dose coverage levels for a new variant that emerges from the wild type cluster on three different dates. Booster dose coverage refers to the percentage of already vaccinated individuals who receive a booster dose. NS refers to values that were not statistically significant from the reference case at a p-value of 0.05.

We next consider how the trade-off between primary series and booster dose coverage changes based upon how well matched the vaccine is antigenically to the emerging variant of concern, now fixing the date of the new variant emergence and varying its level of immune escape (comparing a variant that evades 50% of vaccine and prior ancestral cluster NAbs to one that evades 98% of all prior NAbs). Results suggest that vaccination will avert a smaller percentage of deaths (maximally 21% compared to 62% for a more antigenically similar variant) and will be less efficient overall (nearly 2-fold more doses per death averted) against a variant that evades nearly all vaccine-immunity. However, there is more value in increasing primary series vaccine coverage in terms of percentage of deaths averted, with an up to 7-fold increase in percentage of deaths averted compared to 3.1-fold increase for a booster dose coverage increase, relative to no additional increase in vaccine coverage beyond what has already been achieved) (see Fig 4).

**Fig. 4.**
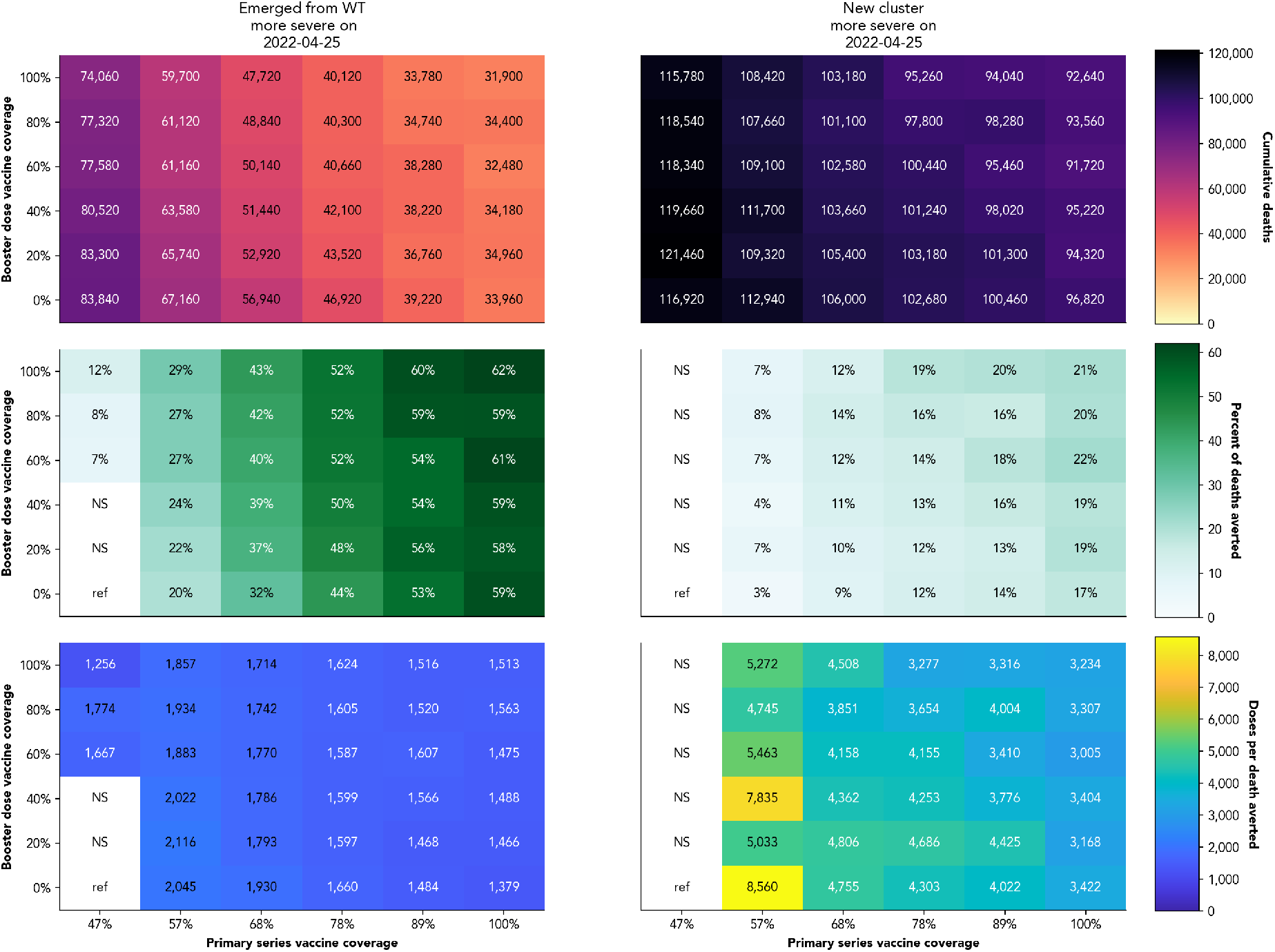
Vaccine impact and efficiency across primary series and booster-dose coverage levels for variants of different antigenic distance from vaccination introduced 6 months following Omicron averaged across 50 replicate simulations. Booster dose coverage refers to the percentage of already vaccinated individuals who receive a booster dose. NS refers to values that were not statistically significant from the reference case at a p-value of 0.05.

### 2.4 Timing of a variant-chasing vaccine strategy

The results from the previous sections suggest that the impact and efficiency of vaccination is highly dependant on the timing and antigenic distance between a renewed vaccine effort and the emerging variant. In light of these findings, we next explored whether a variant-chasing vaccine strategy is effective. We defined a variant-chasing vaccine as the rapid adaptation of our existing vaccines to variant-ready vaccines and boosters that target the specific variant.

The results in Fig 5 reveal that a variant-chasing vaccine strategy has rapidly declining efficacy and efficiency the longer it takes to deploy a variant-specific vaccine following variant introduction. Assuming all vaccinated individuals receive a variant-specific booster series and all unvaccinated individuals receive a variant-specific primary series, a variant-chasing vaccine strategy could avert over 40% of deaths if it is rolled out on a large scale within 5 days from introduction of the new variant, and has a 20-day window in which it would avert more deaths than the same coverage levels with currently available vaccines. The date of introduction of a new variant into a population is impossible to know, but Omicron’s first case was estimated to occur a month before it was first detected (16; 17). Using this lag-time, if it takes 30 days to identify a new variant, the percentage of deaths a variant-chasing vaccine strategy could avert would fall to below 20%, before considering the time it would take to develop and deploy a vaccine after identification.

**Fig. 5.**
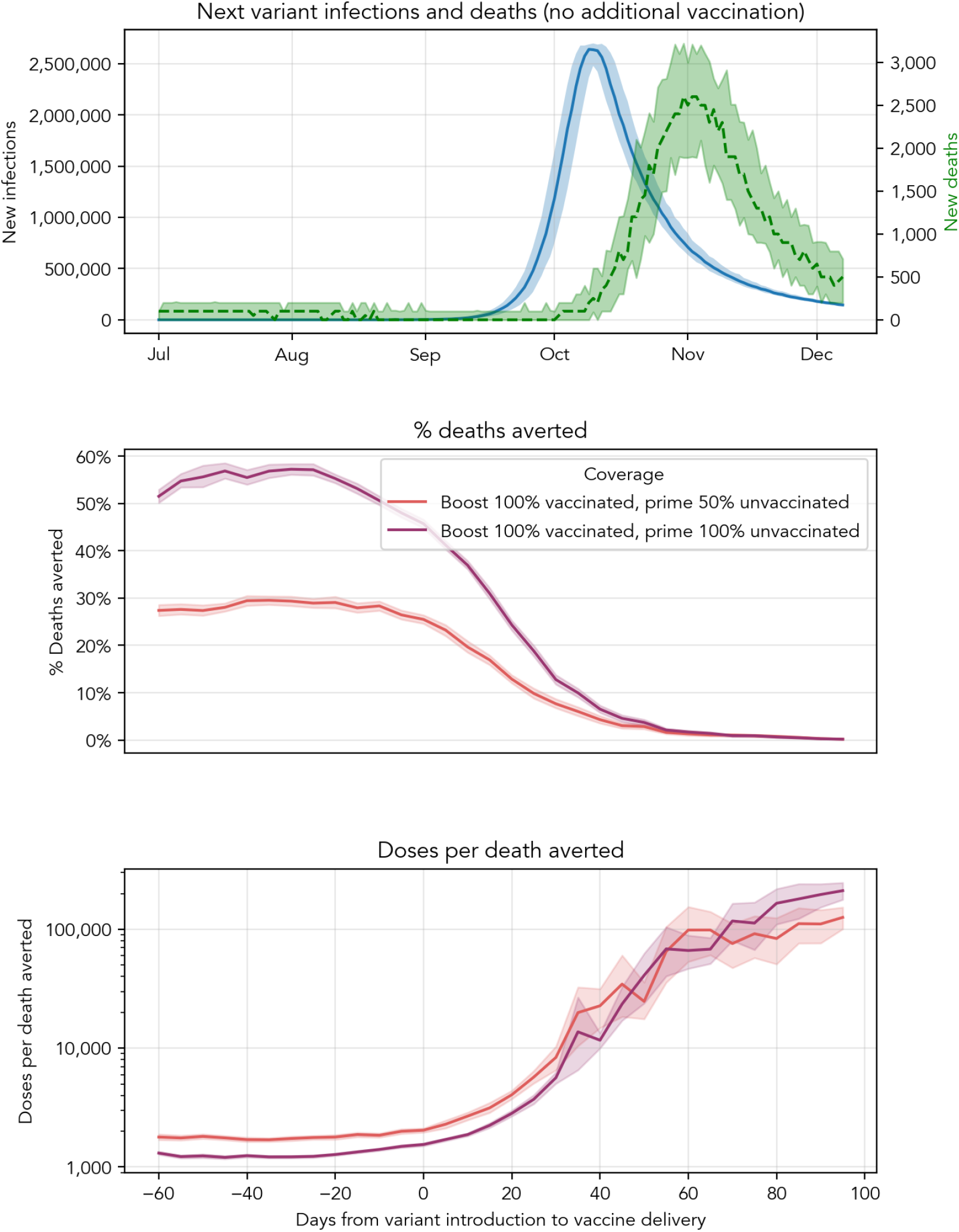
Health impact and efficiency of variant-chasing vaccine. Top panel shows the new infections and deaths associated with the emergence of a new variant 10 months after Omicron that evades 98% of all prior immunity and is 3.5x as virulent as the ancestral strain. The next two panels show percentage of deaths averted and doses per death averted based on how long it takes to begin vaccine roll-out following introduction of a new variant. In these panels, line color indicates what share of already vaccinated and not-yet-vaccinated individuals can be reached with a perfect antigenically-matched vaccine. Negative days imply that the vaccine is rolled out before the variant is introduced, which might occur in a global variant-chasing strategy, where sequencing in the emergent country can inform vaccine delivery in other countries before the variant spreads. All scenarios assume it takes 30 days to reach the target coverage levels once vaccine begins roll-out.

This model has not accounted for geospatial effects that might enable shorter timelines to be possible in some geographies that learn about the emergence of a new variant from other parts of the world. For example, it is estimated that the first case of Omicron in South Africa occurred on approximately October 9, 2021 (16). It was first identified on November 9, 2021 and reported to the WHO two weeks later, after identification of its numerous amino acid changes (17). Two days later it was classified as a variant of concern. By this point, it had spread and approached fixation in South Africa, and so a variant-chasing vaccine in this setting would have minimal impact. However, it might have had a larger impact in other settings which had not yet seen the importation and/or fixation of Omicron, such as the United States, where Omicron was estimated to arrive in late November; Pakistan, India, Brazil, Bangladesh and Mexico in late December; or Indonesia and Russia in middle of January (18). To capture the global utility of variant chasing, we extended the days from variant introduction to vaccine delivery up to two months prior to introduction. One can then reinterpret this figure as representing the possible lag time between variant introduction in the emerging country and vaccine roll-out in a second country. The results suggest that there is approximately a five-week window where developing and deploying a variant-specific vaccine could avert over 50% of deaths, or a 2.5-fold higher impact than vaccinating the same share of the population with the current vaccine. After this time, the benefit begins to fall the longer it takes a vaccine to be deployed. A variant-chasing strategy would avert more deaths than current vaccines at the same coverage levels until 20 days after the introduction of the new variant into a population.

### 2.5 Next generation vaccines

Given the falling efficacy of vaccines over time in common populations with complex immune histories and in the face of immune evading variants, and the challenge of a variant-chasing vaccine strategy, we consider the impact of next generation vaccines that provide broader and more durable (i.e. more slowly waning) immune protection. A vaccine that elicits antibodies that can bind to many diverse strains could protect against highly mutable pathogens, such as SARS-CoV-2 (19). Additionally, a vaccine that provides more durable protection is important in the absence of a seasonal and predictable virus.

Results show that breadth is more important than durability; delivering a broadly neutralizing vaccine could avert up to 40% of deaths, whereas a more durable vaccine alone could maximally avert just over 10% of deaths from a highly immune evading, virulent variant (see Fig 6). However, the impact of breadth alone would fall if the timing of delivery was poorly matched to the timing of the next variant. A durable and broadly neutralizing vaccine would overcome the importance of perfectly timing the vaccine and variant-chasing and could avert up to 65% of deaths in the hypothetical example, an over 3-fold higher impact than vaccinating the same share of the population with the current vaccine against a highly immune-evading variant.

**Fig. 6.**
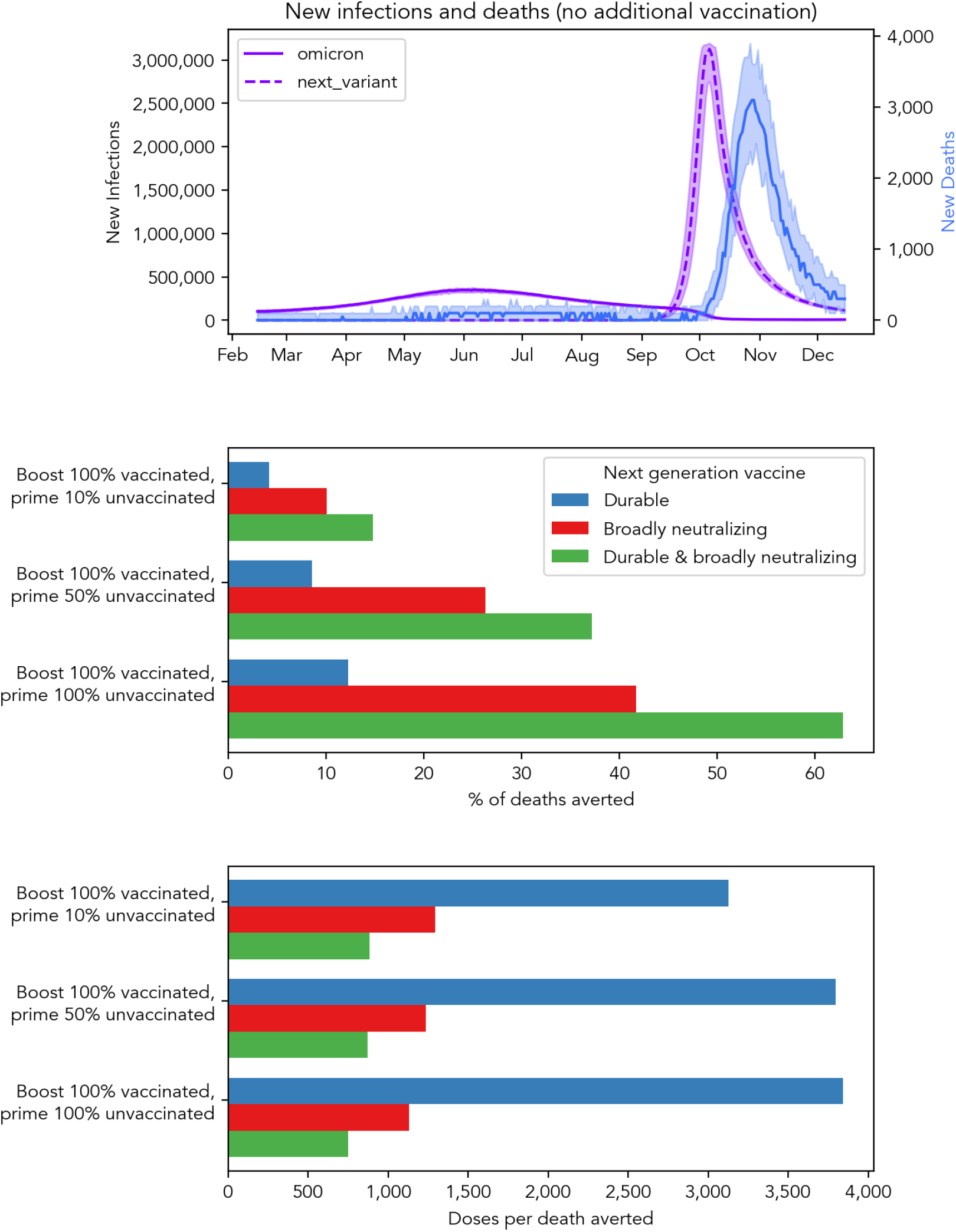
Health impact and efficiency of next-generation vaccines that are more durable and/or more broadly neutralizing. The top panel shows infections and deaths in the absence of additional vaccination. The bottom two panels show the percentage of deaths averted and doses per death averted associated with combinations of next generation vaccines and different levels of target coverage among vaccinated and unvaccinated individuals. Vaccination in these scenarios starts on February 15, 2022 and would take 90 days to reach target coverage levels.

## 3 Discussion

In this work, we used an established agent-based model, Covasim, extended to model intra-host immunity, to explore the changing impact of vaccines over time and in the context of emerging immune evading variants. Our results suggest that the impact of vaccination is highest if delivered in the month prior to an emerging epidemic wave and declines rapidly in its aftermath, and that population vaccine efficiency, in terms of doses per death averted, falls exponentially over time. We show that increasing vaccination coverage among the unvaccinated has a greater marginal and absolute health impact than increasing booster dose coverage among already vaccinated individuals, but that the impact and efficiency of vaccination is highly dependant on the timing and antigenic distance between the vaccine start and the emerging variant. When we consider the implementation of variant-chasing vaccines, we find that while this strategy may never be feasible in the countries in which a variant emerges, it may be a globally effective solution, especially if paired with temporary non-pharmaceutical interventions and improved surveillance that could cut the detection time and possibly slow spread through local restrictions. Alternatively, next generation vaccines that are broadly neutralizing and durable might be necessary as we move into the next phase of COVID endemicity.

SARS-CoV-2 has proven to be a highly adaptable virus and it is nearly certain that evolution will continue and new variants will emerge (20; 21; 22). However, assuming we reach a more stable equilibrium where variants emerge in a seasonal and/or predictable pattern, we may find some of the tools explored here provide options to prevent global spread and burden.

Our results have revealed that a vaccine-only approach has a health impact ceiling. Like flu, a realistic pathway to improving breadth may be vaccinating with variant-specific vaccines to generate broader immunity to now-relevant strains, even if these vaccines are late to stop first waves. Modeling of this approach can be a useful tool for assessing individual- and population-level impact and cost-effectiveness as these vaccines are developed. Additionally, oral antiviral pills may provide a supplemental stopgap measure for reducing disease severity upon emergence of a new variant. This strategy is more agnostic to timing than vaccination, because it can be delivered within 5 days of symptom onset to achieve the full benefit (13). However, while they may be subject to less selection pressure than vaccines, antivirals may lose efficacy over time as well, so will need to be continually re-evaluated as a strategy for pandemic control.

In the aftermath of mass vaccination or infection, it is common to observe a transient phase of low incidence that gives way to recurrent oscillatory dynamics (23; 24). During this period, it is more challenging to measure vaccine efficacy directly, as establishing this requires a prevalent disease (25). It is in situations like this that computer simulation studies, informed by correlates of protection, can be particularly useful (26). A key result of our work is that while vaccine efficacy declines in the wake of a large outbreak, this must be understood as a temporary effect, a corollary of the transient period in which population immunity levels are high. In the absence of ongoing vaccination, both waning immunity and vital dynamics will increase the fraction of the population susceptible to infection, until there are enough for a resurgence (27; 28). Vaccines provide a safe mechanism for counteracting this and preventing or blunting the effect of future resurgences.

Our work demonstrates that the population immunity acquired over the first two years of the pandemic significantly reduces the impact per dose of future vaccinations. In many settings, such as the South-Africa-like one considered here, this immunity is primarily a consequence of what Farmer et al. termed structural violence (29) and has been earned at great loss of life and health. After so much harm has been done, intervention strategies that focus only on delivering vaccines now will further compound that violence, as they provide cover for not addressing other social determinants of health for which no immunity accumulates. While current and next generation vaccines to fight future variants remain an essential part of any effective and equitable strategy, vaccine-based strategies alone are not sufficient. A layered approach to respiratory disease transmission and the social conditions that exacerbate it is required to gain control of this pandemic and prevent the next.

### 3.1 Limitations of the study

Our modeling makes many assumptions that may limit the generalizability of our findings. We do not explicitly characterize co-morbidities or other factors such as immuno-suppression at the individual-level in the model. For these populations who are at highest risk of severe outcomes if infected with SARS-CoV-2, ongoing boosters remain a highly relevant and valuable strategy. We are also not capturing new birth cohorts into the model with near complete susceptibilty and no prior immunity to COVID beyond mother-to-child immune transference, for whom any vaccination strategy would be better than risking infection (30; 31). While our model has a robust mechanistic representation of immune dynamics, we do not capture the process of affinity maturation that antibodies go through over time and after repeated exposures to an antigen that increase the breadth of the immune response even in the face of waning (32). As a result, we may at times be under-estimating the protection retained over time against infection.

## 4 Methods

### 4.1 Model overview

Covasim is an open-source agent-based model developed by the Institute for Disease Modeling with source code and documentation available at https://covasim.org. Covasim simulates individuals interacting via population networks over time, and tracks disease transmission and progression as well as the effects of interventions including symptomatic and asymptomatic testing, isolation, contact tracing, and quarantine, as well as other non-pharmaceutical interventions (NPIs) such as physical distancing, hygiene measures, and protective equipment such as masks. A comprehensive overview of the methodology underlying the model is provided in (33).

Individuals who contract SARS-CoV-2 for the first time then progress through the stages of infection: exposed, infectious (asymptomatic, presymptomatic, mild, severe, or critical), before either recovering or dying, with the probabilities of disease progression dependent on age. Individuals are allowed to recover from any of the disease stages, but only critically ill individuals have a non-zero probability of dying. The model includes individual heterogeneity in infectiousness and in the time spent in each disease state.

Recovered individuals are assumed to be susceptible to reinfection, but their relative susceptibility is modified by a factor that reflects the degree of protective immunity afforded by their prior infection; details on how this factor is calculated are provided in Supplemental Appendix.

As well as belonging to these disease states, agents in the model also have individual attributes that govern their movement through the model over time, such as age, infection history, vaccination history, and whether they have been tested, diagnosed, or contact-traced.

Agents within the model interact with each other within contact networks, which typically stratify contacts in household, workplace, school, and community layers, with the probability of transmission and the efficacy of isolation and quarantine all varying by layer.

The model draws an initial level of neutralizing antibodies (NAbs) following infection or vaccination, tracks the history and source of immunity, captures the priming and boosting of NAbs, and their waning over time (34; 35; 36). At each challenge, “effective NAbs” (Eq 1) is calculated for each individual to determine how much protection is afforded against infection (Eq 5), symptomatic (Eq 6) and severe disease (Eq 7), accounting for cross-immune protection (37).

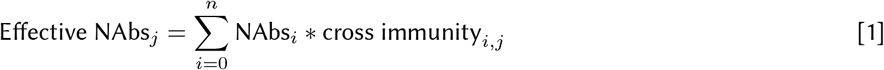

### 4.2 Epidemic input data and model calibration

Our analysis is set in a South African-like setting, meaning we model the age demographics, population size, contact network pattern, and historical immunity seen in South Africa. We collated data on cases, hospitalizations, deaths, and seroprevalence, and combined the available genomic surveillance information to infer the likely dates of introduction of different variants and the background immunity at the seeding time. Serological studies estimated that 19% and 73% of the population had antibodies to COVID-19 in January 2021 and December 2021 respectively (38). In the first part of this analysis, we randomly introduced wild-type, Beta, Delta, and Omicron infections into the population respectively on March 01, 2020, October 15, 2020, May 01, 2021, and October 25, 2021 with variant-specific characteristics as described in Table 1. We modeled the transmission probabilities before and after the four waves of infection to represent changes in non-pharmaceutical interventions that would roughly recreate the levels of seropositivity over time seen in the above-cited studies (39; 40; 38).

**Table 1.** Variant characteristics. Infectivity and severity values refer to the change in per-contact transmission rate and per-infection probability of developing severe disease compared to wild-type. Cross-immunity refers to how much prior neutralizing antibodies are retained to effectively protect against each variant.

| Variant | Infectivity | Severity | WT cross-immunity | Beta cross-immunity | Delta cross-immunity | Pfizer cross-immunity |
| --- | --- | --- | --- | --- | --- | --- |
| Beta | 1 | 3.6 | 6.6% | 100% | 8.6% | 10% |
| Delta | 2.2 | 3.2 | 37.4% | 8.6% | 100% | 33% |
| Omicron | 3.0 | 0.8 | 5% | 4% | 4% | 2.5% |

### 4.3 Vaccine efficacy over time

At the time of writing, South Africa had achieved approximately 47% vaccine coverage of the adult population, achieved over a nine month period, starting with an age- and risk group prioritization, expanding to all adults, and most recently opening up vaccination to everyone over 12 (https://sacoronavirus.co.za/latest-vaccine-statistics/). Sweeping over days from the start of the pandemic through May of 2022, we vaccinate a random 50% of the eligible population (over 18 years of age) with two doses of the Pfizer vaccine and simulate a vaccine trial, randomly selecting participants who are vaccinated on that day as the vaccine arm and participants with no vaccine as the placebo arm. We note that individuals with prior infection are not excluded from either arm of the study, meaning that both the vaccine and placebo arm will be tainted by changing levels of prior immunity over time. We then calculate vaccine efficacy against infection, symptomatic disease, and severe disease over the 60-days following the second vaccine dose.

### 4.4 Vaccine impact in face of new variants

To evaluate the impact of vaccine in the face of new variants, we propose six hypothetical variants that vary in terms of their antigenic distance from existing sources of immunity (41; 42) and their virulence (see Table 2). We chose 3.5-fold more virulent than wild-type as an upper bound of virulence based upon the estimated virulence of the Delta variant. In the absence of a seasonal or known pattern of variant emergence, we randomly introduce the new variant on three dates (4, 6, and 10 months after the introduction of the last variant), representing shorter, equivalent, or longer intervals between variants that have been seen over the last year.

**Table 2.**
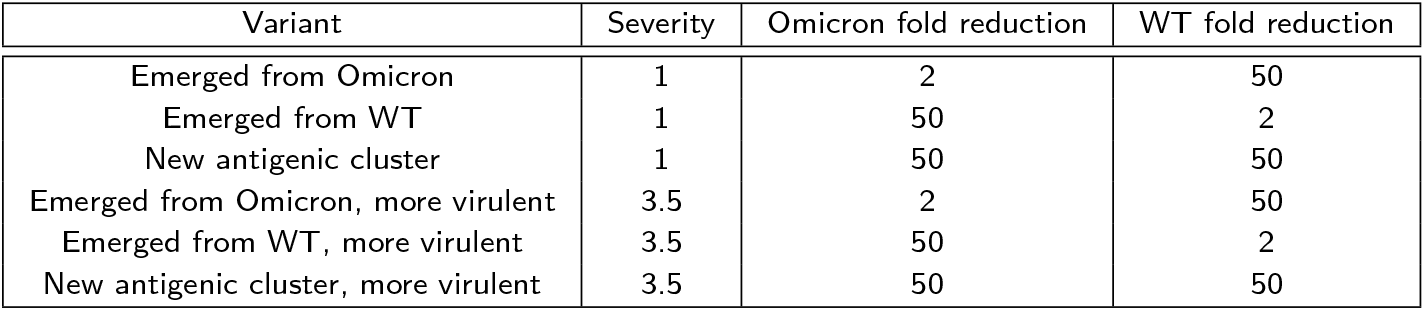
New variant characteristics. Severity values refer to the change compared to wild-type and fold reduction refers to change in neutralization with each variant relative to Omicron or all other (wild-type cluster, including prior variants and vaccines). All new variants were assumed to be 3.5x as transmissible per contact compared to wild-type.

| Variant | Severity | Omicron fold reduction | WT fold reduction |
| --- | --- | --- | --- |
| Emerged from Omicron | 1 | 2 | 50 |
| Emerged from WT | 1 | 50 | 2 |
| New antigenic cluster | 1 | 50 | 50 |
| Emerged from Omicron, more virulent | 3.5 | 2 | 50 |
| Emerged from WT, more virulent | 3.5 | 50 | 2 |
| New antigenic cluster, more virulent | 3.5 | 50 | 50 |

### 4.5 Trade-off between primary series and booster dose coverage

To evaluate the trade-off between primary and booster dose coverage, we simulate the roll-out of vaccine that has been achieved in South Africa to date, starting in May 2021 and reaching 47% coverage of adults over age 18 with two doses by February 2022. We then sweep over a combination of primary series and booster series coverage levels, from 0 to 100% of the eligible population. Eligibility is defined as people over 12 years of age (matching expanded age of vaccination) and unvaccinated for the primary series and over 12 years of age and vaccinated at least 6 months prior for the booster dose. Both the primary and booster-dose series take three months to achieve the target coverage level. We quantify cumulative infections, severe cases, and deaths averted compared to no vaccination for the next 10 months and assuming no further increase in vaccination over this period.

### 4.6 Timing of a variant-chasing vaccine strategy

To evaluate the impact of a variant-chasing vaccine strategy, we consider a variant that emerges to form a new antigenic cluster entirely, meaning it escapes 98% of all prior sources of immunity. A perfectly matched vaccine is rolled out on a set of dates following introduction of the variant. We assume it takes 30 days to reach the target coverage from the date of vaccine roll-out and that the variant-specific vaccine requires two doses to achieve the target efficacy, which we define as the same level of neutralizing antibody as the two-dose primary Pfizer vaccine series.

### 4.7 Next generation vaccines

We explore next generation vaccines in the model by comparing the impact of vaccines with combinations of these properties delivered today against an emergent immune evading variant 10 months following Omicron. We specifically define a more broadly neutralizing vaccine as having complete cross-immunity against future variants and a more durable vaccine as waning at a significantly slower rate than current vaccines. Considering combinations of vaccines with these properties, we model three vaccine coverage scenarios that vary what share of currently unvaccinated individuals would be reached with the next generation vaccines and assume all currently vaccinated individuals would receive the vaccine once eligible (eligibility defined as 6 months following second dose).

A fully reproducible repository can be found at https://github.com/amath-idm/post-omicron.

## Data Availability

All data produced in the present study are available upon reasonable request to the authors.

https://github.com/amath-idm/post-omicron

## 5 Acknowledgments

The findings, conclusions, and views expressed in this study are those of the author(s) and do not necessarily represent those of the Bill & Melinda Gates Foundation.

## 6 Author Contributions

JAC, DJK, MF conceived of the study. The model was developed by JAC, CCK, RMS, and RA. JAC programmed and analyzed the model, produced visualizations, and drafted the manuscript. All authors reviewed and edited the manuscript. Supervision was provided by DJK and MF.

## A Supplementary Material

### A.1 Quantifying neutralizing antibodies as a correlate of protection

Work from Khoury et al. (34) and Earle et al. (43) relates neutralizing antibodies to vaccine efficacy, showing that neutralization level is highly predictive of immune protection from symptomatic COVID-19, and that despite decaying immunity, protection from severe disease should be largely retained. However, this work did not consider immune protection from infection and did not disentangle the relationship between NAbs and protection against primary infection, symptomatic, and severe disease.

We model protective efficacy as a function of neutralizing antibodies (NAbs), informed by data from vaccine immunogenicity and efficacy trials as well as observational studies. Extending upon the methodology from Earle (43) and Khoury (34), we estimate efficacy for primary SARS-CoV-2 infection, symptomatic COVID-19, and severe disease jointly and calculate conditional efficacies for symptom- and severity-blocking given infection, revealing a direct model of NAbs as a correlate of protection. We additionally consider differences between the neutralizing antibodies generated from vaccines and infection while accounting for antibody waning between immunogenicity and vaccine efficacy studies.

#### A.1.1 Data extraction

In order to map neutralizing antibody (NAb) level to protective efficacy, we extracted cohort estimates from vaccine immunogenicity and efficacy trials as well as data on reinfection. In the absence of standardized assays to measure NAbs, normalization against a convalescent serum standard has been suggested as a method for providing greater comparability between results from different assays (44). In order to compare the immunogenicity data with the efficacy endpoints, we accounted for waning that may have occurred across the timescales reported. We re-normalize the average NAb for each of the cohorts using an adaptation of the antibody kinetics functional form described in Khoury et al. (34) fit to cohorts of hospitalized patients and healthcare workers followed-up for eleven months after COVID-19 symptom onset (45).

In the NAb re-normalization procedure, we used a model of immune waning to account for any decay in antibodies that may have occured between the time of the antibody assay collection and vaccine efficacy endpoints. To do so, we assume waning follows a 2-part exponential decay and fit the half-life and duration parameters to cohorts of French and Irish hospitalized patients and healthcare workers followed for up to eleven months after COVID-19 symptom onset (45). Relative to the waning model used by Khoury et al., our model suggests both shorter initial decay of NAbs followed by a steeper long-term decay rate.

We also adjusted the reported neutralization level in settings where variants of concern were circulating at the time of efficacy endpoints based upon reported neutralization in convalescent and vaccine sera. In order to compute titer shifts for efficacy against variants of concern, NAbs were randomly shifted by a normally-distributed scaling factor,

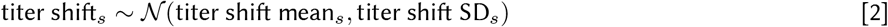

with a mean and standard deviation based upon titer shifts reported in the literature, see Table 3.

**Table 3.** Variant neutralization with wild-type and vaccine-sera. Values refer to the fold of reduction in neutralization with each variant relative to wild-type infection or vaccine source, standard deviation reported in parentheses.

| Variant | Wild-Type | Pfizer | Moderna | AstraZeneca |
| --- | --- | --- | --- | --- |
| Alpha (B.1.1.7) | 1.8 (0.41) | 1.8 (0.41) | 1.8 (0.41) | – |
| Beta (B.1.351) | 13.3 (2.17) | 10.3 (2.9) | 12.4 (2.85) | – |
| Gamma (P.1) | 8.66 (1.05) | – | – | – |
| Delta (B.1.617.2) | – | 2.19 (0.05) | – | 4.01 (0.32) |

#### A.1.2 Immune model

We modeled three types of immunity: protection against infection, symptomatic disease, and severe disease and fit separate functions for immunity derived from infection and vaccination, which can be supported by the role of nucleocapsid-specific antibodies which are missing from some vaccines and may mechanistically explain why infection NAbs are more effective against infection (46).

We jointly estimated the relationship between NAbs and protective efficacy against infection, symptoms, and severe disease with study-specific random effects. VE_symp|inf,*r*_ and VE_sev|symp,*r*_ are unobserved, and we model them through the marginal efficacy against symptomatic and severe disease (Eqs 3 and 4).

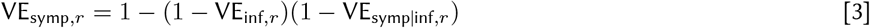

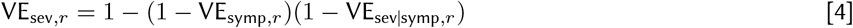

We split vaccine efficacy into conditional parts to match the stages of the infection process and assumed both the efficacy against infection and the conditional efficacy against symptoms and severe disease are logit-log. The *α* and *β* parameters capture the intercept and slope in each equation, respectively (see Eqs 5, 6, 7). Vaccine efficacy against infection, VE_inf_, is the first stage that modulates the probability of infection given exposure. For people who get infected, symptomaticity is modulated by the conditional vaccine efficacy given breakthrough infection, VE_symp|inf_, and similarly severity is modulated by the conditional vaccine efficacy given a breakthrough symptomatic infection VE_sev|symp_.

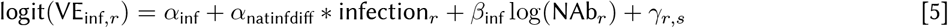

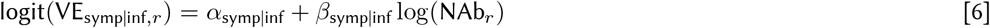

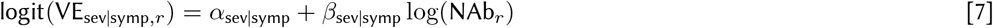

where NAb_*r*_ represents the average level of neutralizing antibodies across participants in record *r*, “infection” is a dummy variable that is equal to 1 when record *r* is immunity from infection and equal to 0 when record *r* is immunity from vaccination, and *γ*_r,s_ is the random effect from study *s* associated with record *r* (some studies have multiple associated records).

We assumed study random effects are normally distribution with a mean of 0 and a standard deviation that is Cauchy distributed with a flat prior,

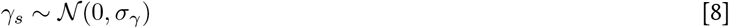

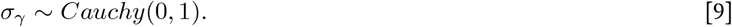

For studies which reported efficacy against variants of concern, as part of the re-normalizing computational procedure, NAbs were randomly shifted by a normally-distributed scaling factor with a mean and standard deviation based upon titer shifts reported in the literature (47; 48; 49; 50; 51; 51; 52). That is, the results marginalize over uncertainty in the NAb titer shift.

We estimated a Bayesian posterior for parameters in Eqs 5 - 7 with a Hamiltonian Monte Carlo method fit in Stan based upon the likelihood (Eqs 10 - 12). 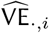 denotes vaccine efficacy estimators arising from the studies, that is, the data. The likelihood uses the standard errors as reported by the studies adapted to a log-scale as a reasonable proxy for the true standard deviation, one that incorporates study sample size.

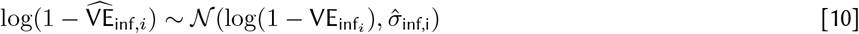

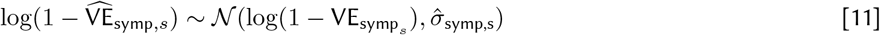

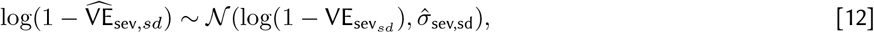

for *k* in *K* unique study cohorts, *i* in *I* infection records, *s* in *S* symptomatic disease records, and *sd* in *SD* severe disease records.

In the above equations, *σ*_*γ*_ is the standard deviation of the study random effects, *γ*_*s*_ represents the study-specific random effect *s* in *S* unique studies, and 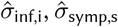, and 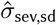 represent the log-scale standard deviation of outcomes of protection against infection, symptomatic and severe disease.

Model fitting was performed in R using Stan.

#### A.1.3 NAbs as a correlate of protection

We find that neutralizing antibodies are strongly correlated with protection against SARS-CoV-2, symptomatic COVID-19, and severe disease (see Fig 7). In order to provide a 50 percent or higher reduction in the risk of symptomatic COVID-19, a vaccine would need to induce a NAb level at least one-tenth of the average convalescent level, and a one-third NAb level would be required to reduce the risk of infection by 50 percent or higher. While natural infection provides greater protection than vaccination for the same level of NAbs, all of the vaccines considered in this analysis meet the 50 percent risk reduction threshold and do not have the morbidity and mortality costs associated with COVID-19 infection. However, as will be discussed below, variants of concern challenge the efficacy of vaccines by reducing the neutralization levels and associated protection.

**Fig. 7.**
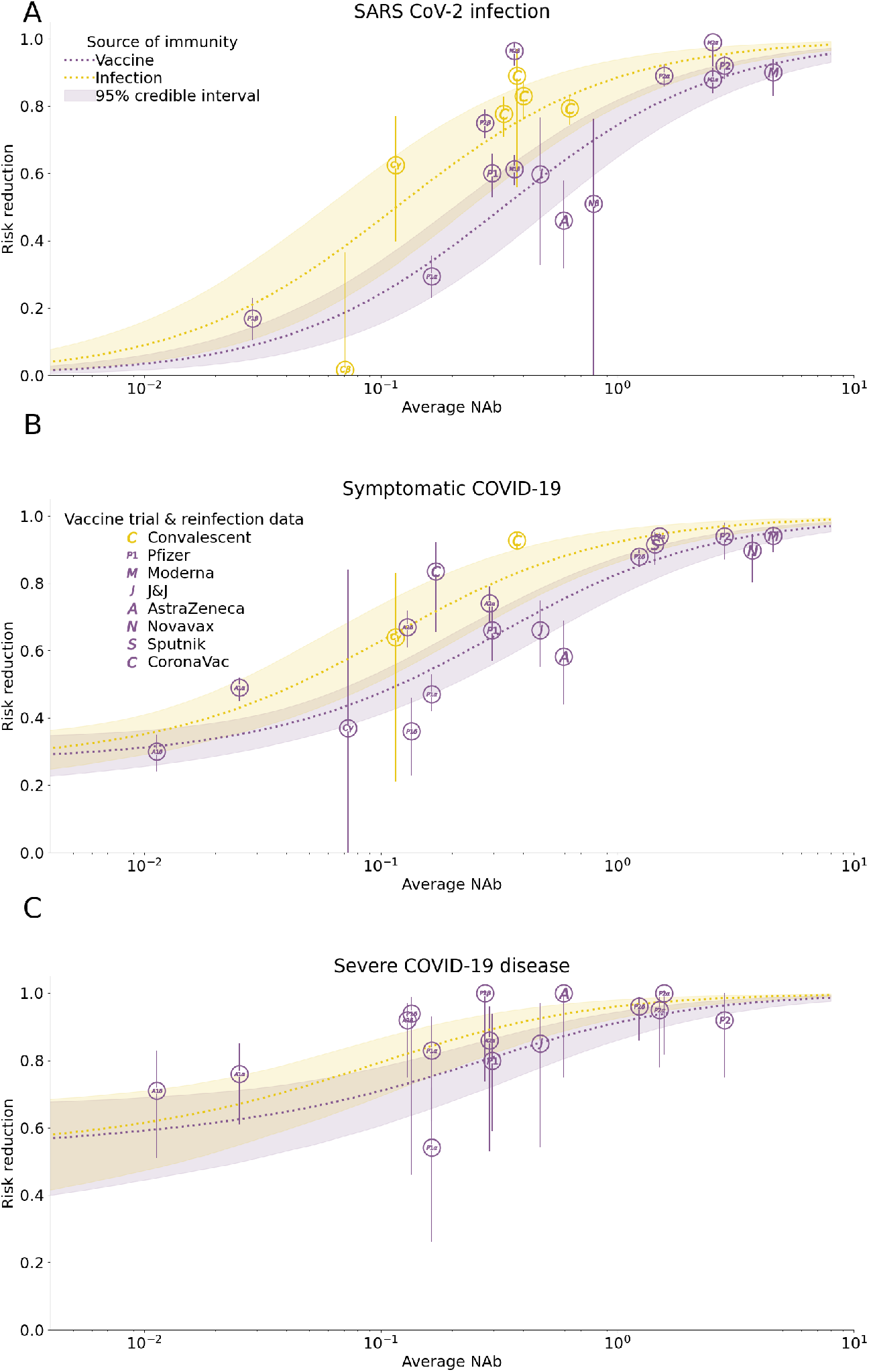
Immunity model. We jointly estimated the relationship between NAbs and vaccine efficacy against infection, symptomatic, and severe disease using meta-study data. NAbs are normalized relative to human convalescent sera and central estimates are plotted. Panels show fitted efficacy against (A) infection, (B) symptomatic COVID-19, and (C) severe COVID-19 as a function of infection-and vaccine-derived NAbs. Shaded regions represent 95% credible intervals with *γ*_*k*_ fixed at its median.

Given the fitted marginal efficacies above, we inferred the conditional protection against symptomatic and severe disease for individuals with a breakthrough infection and with breakthrough symptomatic disease. We find that any history of immunity would provide some protection against symptomatic and severe COVID-19, with a floor of approximately 37 percent reduction in the risk of symptomatic COVID-19 and 50 percent reduction in risk of severe disease conditional on a breakthrough infection or disease respectively (see Fig 8). From that level, a percentage increase in NAbs would result in 3.8 percent (0.12, 11.75) reduction in risk against symptomatic disease a 7.9 percent (0.22, 26.15) reduction in the risk of severe COVID-19, conditional on a breakthrough infection and breakthrough disease respectively. While NAbs are correlated with protection against COVID-19, there may be other immune mechanisms, such as T-cell response, that provide protection against symptomatic and severe disease (53). These results suggest that even as antibodies wane and become insufficient to protect against infection, some immunity to symptomatic and severe disease will remain.

**Fig. 8.**
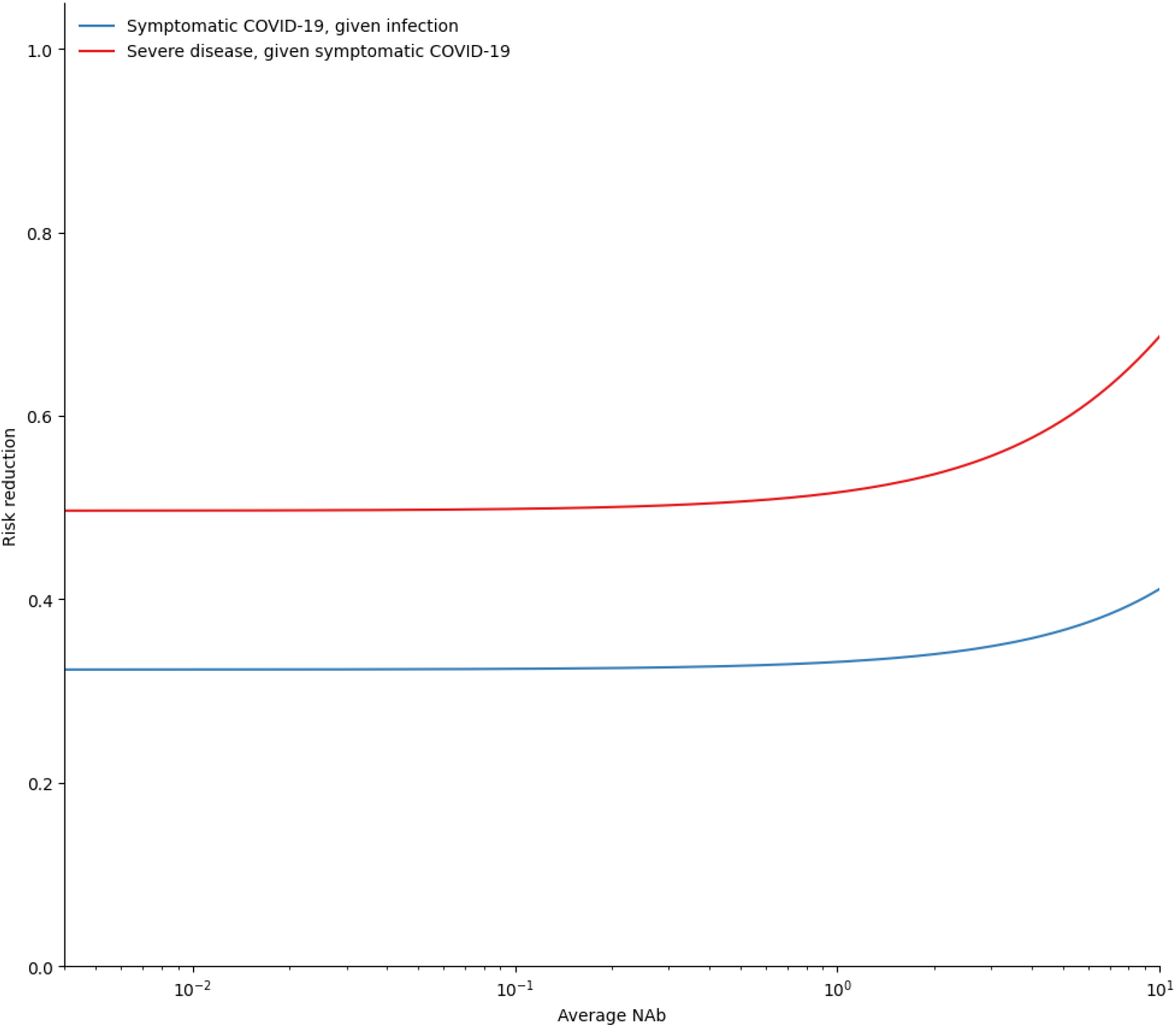
Conditional protection against symptomatic and severe disease given a breakthrough infection and breakthrough symptoms, respectively.

We report the mean and standard deviation of the model parameters in Table 4.

**Table 4.** Fitted parameter values based upon HMC algorithm with 30,000 iterations across 5 chains.

|  | Mean | Standard Deviation |
| --- | --- | --- |
| $\alpha_{inf}$ | 1.08 | 0.226 |
| $\alpha_{natinf diff}$ | 1.0 | 0.403 |
| $\beta_{inf}$ | 0.967 | 0.055 |
| $\alpha_{symp\vert inf}$ | -0.739 | 0.187 |
| $\beta_{symp\vert inf}$ | 0.038 | 0.0317 |
| $\alpha_{sev\vert symp}$ | -0.0143 | 0.311 |
| $\beta_{sev\vert symp}$ | 0.0799 | 0.078 |
| $\sigma_{\gamma}$ | 0.639 | 0.177 |

Our results show that while NAbs induced by natural infection or vaccination wane and individuals may lose sterilizing immune protection, immune memory is likely to be retained long-term to provide significant protection against severe disease, even in the face of immune-evading variants. This suggests that neutralizing titers play a large role in preventing infection, but that other immunologic factors may play a more dominant role in controlling infection once it occurs.

Our modeling approach relies on estimating a relationship between NAbs and protection against infection, symptomatic disease, and severe disease, and the data used to establish these estimates are scarce and uncertain, especially for low levels of NAbs. While a full individual-level model would be ideal, we relied upon published cohort averages and tried to account for variation and heterogeneity between studies using study-level random effects. We also assume that the antibody kinetics are identical for vaccine- and naturally-derived NAbs. As more longitudinal immune studies emerge, we will have the opportunity to test and refine this hypothesis. We do not specifically model cellular immune responses, although they are likely to also influence disease symptomaticity and severity and to have different kinetic profiles than antibodies (53; 54).

## Notes

### Competing Interest Statement

The authors have declared no competing interest.

